# Association between APOE genotypes and Metabolic Syndrome in a Middle Aged and Elderly Urban Indian Population

**DOI:** 10.1101/2024.06.11.24308744

**Authors:** Shilna Azhuvalappil, Raghav Prasad, Pravin Sahadevan, Priya Chatterjee, Hitesh Pradhan, Pooja Rai, Anant Gupta, Reddy Peera Kommaddi, Thomas G. Issac, Jonas S. Sundarakumar

**Affiliations:** Centre for Brain Research, Indian Institute of Science, Bangalore 560012, India; Indian Institute of Science Education and Research (IISER), Kolkata 741246, India; Manipal Academy of Higher Education, Manipal, India

**Keywords:** Apolipoprotein E (APOE), Metabolic Syndrome, Aging, Cardiovascular Diseases, Lipid Metabolism

## Abstract

**Background:** This study examines the association between apolipoprotein E (APOE) genotypes and metabolic syndrome (MetS) in an older urban population in South India, as part of the Tata Longitudinal Study on Aging.

**Methods:** A total of 618 participants aged 45 and above were analyzed cross-sectionally for the association between APOE carrier status and MetS (based on both NCEP ATP III and Consensus criteria).

**Results:** Despite the high prevalence of MetS observed in this cohort (51.62% by NCEP-ATP III and 61.33% by Consensus criteria), multivariable logistic regression revealed no significant association between APOE genotypes and MetS under both criteria. However, specific associations were noted in age and sex-stratified analyses; notably, E2 carriers under 60 showed 0.42-fold decreased odds (95%CI:0.20,0.89, p-value-0.023) for an increased waist circumference, and E4 carriers above 60 were at 1.85 times increased odds (95% CI:1.04,3.28, p-value<0.05) for decreased HDL.

**Conclusion:** These findings suggest that while APOE genotypes influence certain metabolic parameters, their impact on MetS may be limited in this urban setting, possibly overshadowed by environmental factors and lifestyle influences which was highlighted by the differences seen in its sister rural cohort.

## Introduction

Metabolic syndrome (MetS) is the concurrent presence of elevated blood pressure, hyperglycemia, dyslipidemia, and abdominal obesity (1). MetS has been associated with a two-fold increased risk for cardiovascular disease (CVD) and a 1.5-fold increased risk of all-cause mortality (2).

APOE plays a crucial role in managing various blood lipids, affecting the levels of triglycerides (TG), and lipoprotein particles in the plasma (3). The APOE gene has three common alleles—ε2, ε3, and ε4—which produce three major isoforms: E2, E3, and E4. These alleles result in three homozygous genotypes (E2/2, E3/3, E4/4) and three heterozygous genotypes (E2/3, E2/4, E3/4). Each allele has a distinct binding affinity for lipoprotein receptors, which significantly impacts lipid transport, thus contributing to differing risks for MetS (4). Studies in diverse populations have noted associations between the APOE E4 allele and metabolic syndrome. However, data from the Indian populations on this relationship are scarce.

Our study examines the link between APOE genotypes and metabolic syndrome (MetS) and its components in middle-aged and older adults from urban India. We then compare these findings with those from a harmonized rural sister-cohort (5). Furthermore, considering previous findings that the effects of APOE vary with age and sex, particularly in factors like hypertension (6,7), we will also employ an age and sex-stratified approach. We hypothesize that APOE carrier status is associated with MetS.

## Methods

The present study is a cross-sectional analysis of data from an ongoing aging cohort study in urban southern India named the Tata Longitudinal Study on Aging (TLSA), which recruits participants aged 45+ years from the city of Bangalore, India.

The analytical sample for this study (n=618) includes participants who had undergone their baseline clinical and biochemical assessments between 2015, June, and 2023, December and had complete data on the variables included in this study. Individuals diagnosed with dementia, severe medical or psychiatric illnesses, and sensory or motor impairments that could affect the study evaluations were excluded. Further, participants with APOE □2/□4 genotype were excluded due to the potentially contrasting effects of □2 and □4 alleles on lipid levels.

Clinical and anthropometric measures were collected using a structured clinical questionnaire that gathered demographic characteristics, clinical history of comorbidities, and data on physical inactivity, tobacco, and alcohol consumption.

Anthropometric data, including waist circumference (WC) and blood pressure, were measured by trained nurses. Biochemical investigations involved fasting peripheral venous blood samples analyzed by an accredited lab for glucose, triglycerides, LDL, and HDL.

Metabolic syndrome (MetS) was defined according to the NCEP ATP III (8) and Consensus criteria (9). APOE genotyping methods are detailed elsewhere (10). Participants were classified into E2 carriers (with either □2/□2 or □2/□3 genotype), E4 carriers (with either □4/□4 or □3/□4 genotype), and E3 homozygous (□3/□3 genotype) individuals, who served as the reference group in the logistic regression model.

Participant characteristics were summarized as frequencies and percentages for categorical variables and means with standard deviations for continuous ones. We tested differences between variables and metabolic syndrome (MetS), defined by NCEP ATP III criteria, using independent two-sample t-tests for continuous variables and chi-square tests for categorical variables. Multivariable binary logistic regression assessed the association between APOE genotypes and MetS and its components, adjusting for covariates. A p-value of <0.05 was deemed statistically significant. All analyses were conducted using Stata version 18.0.

## Results

Out of the 618 study participants, 319 (51.62%) were diagnosed with MetS, respectively (as per the NCEP ATP-III criteria). There were significant differences in the distribution of age, sex, education, diabetes mellitus, hypertension, cardiac illness, high blood pressure, hyperglycemia, hypertriglyceridemia, low HDL-C, body mass index, and high waist circumference between the MetS and no Mets group **(Table 1)**. There was no statistically significant difference in the distribution of APOE genotypes by MetS in both criteria **(Table 1 & Supplementary Table 1)**.

**Table 1.**
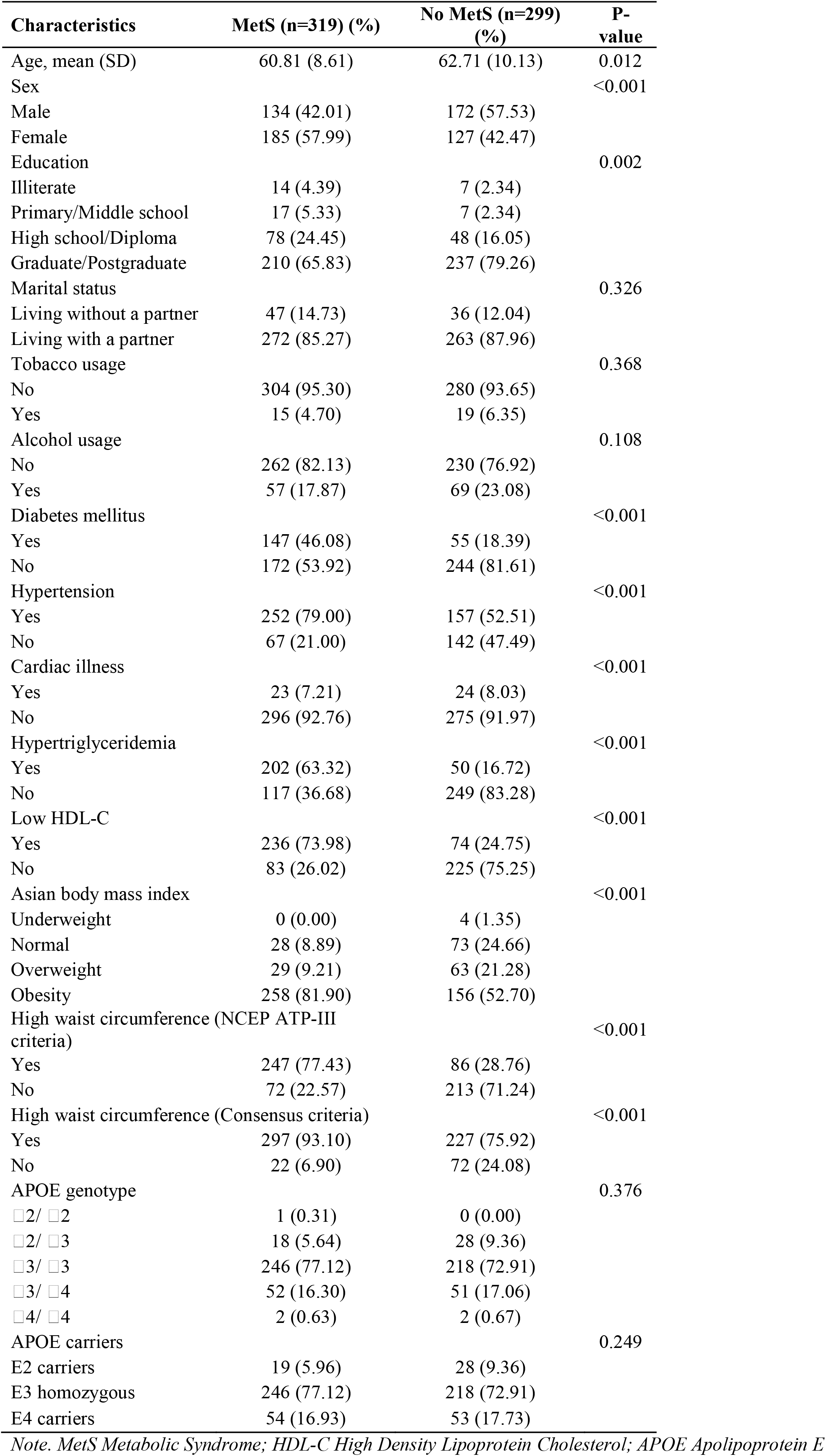
*Demographic, anthropometric, and plasma biochemical characteristics of the participants of rural cohort (MetS defined by NCEP ATP-III criteria)*

Multivariable logistic regression models showed no significant association between APOE genotypes and metabolic syndrome per the NCEP ATP III or the Consensus criteria. Among the individual components of MetS, we only found that E2 carriers exhibited 0.42-fold decreased odds for high waist circumferences (95% CI: 0.20,0.89, p-value-0.023) according to the consensus criteria. **(Table 2)**.

**Table 2.** Association between ApoE genotype and individual component of MetS.

| Outcome | APOE3 | APOE2 | APOE4 |
| --- | --- | --- | --- |
| High blood pressure <sup>g</sup> | 1.00<br>(Reference) | 0.71<br>(0.35-1.44) | 0.81<br>(0.50-1.31) |
| Hyperglycemia <sup>g</sup> | 1.00<br>(Reference) | 1.23<br>(0.63-2.40) | 1.08<br>(0.69- 1.70) |
| Hypertriglyceridemia <sup>g</sup> | 1.00<br>(Reference) | 1.34<br>(0.69-2.59) | 1.05<br>(0.66-1.66) |
| Low HDL-C <sup>g</sup> | 1.00<br>(Reference) | 0.84<br>(0.43-1.63) | 1.43<br>(0.91-2.24) |
| High waist circumference (NCEP ATP-III criteria) <sup>g</sup> | 1.00<br>(Reference) | 0.61<br>(0.30-1.23) | 0.89<br>(0.54-1.45) |
| High waist circumference (Consensus criteria) <sup>g</sup> | 1.00<br>(Reference) | **0.42**<br>**(0.20-0.89) \*** | 0.83<br>(0.45-1.54) |
| Mets (NCEP ATP-III criteria) <sup>f</sup> | 1.00<br>(Reference) | 0.68<br>(0.36-1.29) | 0.92<br>(0.59-1.42) |
| Mets (Consensus criteria) <sup>f</sup> | 1.00<br>(Reference) | 0.79<br>(0.43-1.48) | 1.15<br>(0.73-1.80) |

*Note. MetS Metabolic Syndrome; HDL-C High Density Lipoprotein Cholesterol, APOE Apolipoprotein E*
*\*P-value<0.05*
*<sup>f</sup> The logistic regression model adjusted for age, education, marital status, physical activity, alcohol usage, tobacco usage and cardiac illness*
*<sup>g</sup> Each component of Mets was adjusted for other components along with sex, age, marital status, physical activity, alcohol usage, tobacco usage and cardiac illness*

On stratifying the population by sex, we again found no significant association between APOE and MetS for either criterion or its components in both sexes (**Supplementary Table 2)**.

Upon age stratification (<60 and ≥ 60) there was no association seen between MetS and APOE in either group. In the group < 60 years we found that E2 carriers had decreased odds for high WC (OR: 0.18, 95% CI: 0.04,0.88, p-value < 0.05). In the age group of ≥60 years for E4 carriers we found that they had 1.85 times increased odds (95% CI: 1.04,3.28, p-value < 0.05) for decreased HDL (**Supplementary Table 3)**.

## Discussion

Our study of urban Indians aged 45 and older found a high prevalence of metabolic syndrome: 51.62% by NCEP-ATP III criteria and 61.33% by Consensus criteria. Whereas the prevalence in the rural sister cohort was 38.5% and 46.5% respectively. This compares to a national prevalence of 41-50% for a similar age group reported in a recent meta-analysis (11). Even among the participants who did not meet the criteria for metabolic syndrome, there was a high prevalence of its components, ranging from 16% to 54%.

However, we found no significant association between APOE and MetS. Prior studies from various populations have revealed conflicting findings, reporting a significant association (12–14), while others from India (15) and China (1) have reported a lack of association. Such variations in the findings could be attributed to the differences in the demographics of the populations examined, different inclusion and exclusion criteria, and the use of different criteria for diagnosis of MetS.

Recent findings from the SANSCOG study revealed a sex-specific association between APOE and MetS, wherein the rural females had increased odds for MetS, hypertriglyceridemia, and low HDL (10). In contrast, this study found no significant association with most of the MetS components, which is also contrary to the existing literature where APOE is significantly associated with lipid, glycemic (16), and blood pressure parameters (17). However, when we age-stratified, we saw that participants aged 60 years and above had increased odds of low HDL. Previous studies have indicated that the interaction of APOE with components of MetS changes with age (7). Accordingly, we found a significant association between high WC (consensus criteria) and APOE genotype in the total population which on age stratification was only present in the <60 age group.

The discordance of our results with our sister study and the overall contrasting evidence for APOE polymorphisms and MetS could be due to varying exposure to environmental risk factors, which, in turn, could have diminished the genetic influence of APOE on MetS. Further, differential healthcare access and treatment of the metabolic risk factors could potentially confound this association. Finally, since there is tremendous genetic diversity in the Indian population, it is possible that the lack of association between the APOE allele and MetS could be unique to our study population. Significant differences in APOE E4 allele frequency have been reported in the Indian population compared to other populations (18). The role of APOE has been most widely studied as a risk factor for dementia and cognitive performance and differences in the association between APOE allele status and dementia/cognitive performance across ancestries and ethnicities have been reported (19). Linkage disequilibrium, allele frequencies, and genetic architecture of a population could modulate the relationship between genetic effects and diseases (20) .

The limitations of our study include its small sample size, which may hinder the generalizability of our findings. Additionally, the use of convenience sampling might not provide a sample that is representative of the broader urban Indian population. The cross-sectional design of the study also prevents us from establishing causality between APOE genotypes and MetS. Furthermore, we did not consider several other genetic factors that could influence the MetS burden.

To the best of our knowledge, this is the first and most comprehensive study from India to assess the association between APOE and MetS in the middle-aged and older urban population with an objective diagnosis of all MetS components. Also, this paper is the first of its kind to compare APOE polymorphisms between rural and urban populations from two harmonized studies which are within a small geographical distance thus making the comparison reliable. We also looked at sex-specific and age-specific associations and gathered important insights that align with the limited previous literature on the same.

In conclusion, environmental factors in the urban population could potentially outweigh the genetic predisposition in determining health risks, therefore, highlighting the requirement of lifestyle modification in managing cardio-metabolic diseases in this population.

## Data Availability

All data produced in the present study are available upon reasonable request to the authors.

## Ethics Approval Statement

All participants provided written informed consent before their involvement in the study. The study protocol received approval from the Institutional Review Board at the Institute. All experiments conducted adhered to the appropriate guidelines and regulations. The Institutional Ethical Clearance number provided for this study is CBR/42/IEC/2022-23.

## Author Contributions

Conception: JSS, SA; Acquisition, Analysis, or Interpretation of data: JSS, TGI, RPK, PR, SA, PS, HP, AG; Drafting the work or revising: SA, PC, RP, JSS, TGI; Final approval of the manuscript: JSS, TGI, RPK, PC, RP.

## Acknowledgments

We are grateful to the volunteers who participated in the CBR-TLSA study. We acknowledge all members of the CBR-TLSA study team for their valuable contributions to various aspects of the CBR-TLSA study.

