## Supplementary Material for "Association between APOE genotypes and Metabolic Syndrome in a Middle Aged and Elderly Urban Indian Population"

**Supplementary Table 1**

*Distributions of the ApoE genotypes by MetS status based on the Consensus criteria*

| **MetS (Consensus criteria)** | **Mets (n=379) (%)** | **No Mets(n=239) (%)** | **P - value** |
| --- | --- | --- | --- |
| APOE carriers |  |  | 0.460 |
| E2 carriers | 25 (6.60) | 22 (9.21) |  |
| E3 homozygous | 286 (75.46) | 178 (74.48) |  |
| E4 carriers | 68 (17.94) | 39 (16.32) |  |

*Note. Mets- Metabolic Syndrome, APOE- Apolipoprotein E*

**Supplementary Table 2**

*Association between ApoE genotype and individual component of MetS stratified by sex.*

| **Outcome** | **Males** | | | **Females** | | |
| --- | --- | --- | --- | --- | --- | --- |
|  | **APOE3** | **APOE2** | **APOE4** | **APOE3** | **APOE2** | **APOE4** |
| High blood pressure g | 1.00 (Reference) | 0.80 (0.29-2.21) | 0.78 (0.38-1.59) | 1.00 (Reference) | 0.73 (0.26-2.10) | 0.93 (0.46-1.88) |
| Hyperglycemia g | 1.00 (Reference) | 1.24 (0.50-3.07) | 1.01 (0.53-1.91) | 1.00 (Reference) | 1.16 (0.42-3.24) | 1.10 (0.57-2.11) |
| Hypertriglyceridemia g | 1.00 (Reference) | 0.91 (0.34-2.44) | 1.31 (0.57-2.25) | 1.00 (Reference) | 2.09 (0.77-5.65) | 1.22 (0.64-2.33) |
| Low HDL-C g | 1.00 (Reference) | 1.27 (0.51-3.20) | 0.96 (0.49-1.85) | 1.00 (Reference) | 0.64 (0.24-1.75) | 1.97 (1.00-3.88) |
| High waist circumference (NCEP ATP-III criteria) g | 1.00 (Reference) | 0.46 (0.16-1.36) | 0.64 (0.31-1.30) | 1.00 (Reference) | 0.87 (0.32-2.37) | 1.11 (0.51-2.41) |
| High waist circumference (Consensus criteria) g | 1.00 (Reference) | 0.42 (0.17-1.04) | 0.90 (0.43-1.90) | 1.00 (Reference) | 0.31 (0.70-1.34) | 0.45 (0.14-1.51) |
| Mets (NCEP ATP-III criteria) f | 1.00 (Reference) | 0.70 (0.29-1.68) | 0.66 (0.29-1.68) | 1.00 (Reference) | 0.61 (0.24-1.55) | 1.19 (0.63-2.26) |
| Mets (Consensus criteria) f | 1.00 (Reference) | 0.74 (0.31-1.75) | 1.14 (0.60-2.15) | 1.00 (Reference) | 0.84 (0.33-2.13) | 1.14 (0.60-2.18) |
| **P-value<0.05* |  |  |  |  |  |  |
| *f The logistic regression model adjusted for age, education, marital status, physical activity, alcohol usage, tobacco usage and cardiac illness* | | | | | | |
| *g Each component of Mets was adjusted for other components along with gender, age, marital status, physical activity, alcohol usage, tobacco usage and cardiac illness* | | | | | | |

**Supplementary Table 3**

*Association between ApoE genotype and individual component of MetS stratified by age group.*

| **Outcome** | **Below 60** | | | **60 and above** | | |
| --- | --- | --- | --- | --- | --- | --- |
|  | **APOE3** | **APOE2** | **APOE4** | **APOE3** | **APOE2** | **APOE4** |
| High blood pressure g | 1.00 (Reference) | 0.53 (0.12-2.31) | 0.74 (0.33-1.67) | 1.00 (Reference) | 0.87 (0.37-2.01) | 0.85 (0.45-1.61) |
| Hyperglycemia g | 1.00 (Reference) | 1.84 (0.39-8.76) | 0.95 (0.42-2.13) | 1.00 (Reference) | 1.03 (0.49-2.17) | 1.03 (0.59-1.80) |
| Hypertriglyceridemia g | 1.00 (Reference) | 1.40 (0.34-5.84) | 1.79 (0.81-3.92) | 1.00 (Reference) | 1.35 (0.63-2.93) | 0.82 (0.45-1.49) |
| Low HDL-C g | 1.00 (Reference) | 0.37 (0.08-1.71) | 1.10 (0.50-2.41) | 1.00 (Reference) | 1.14 (0.52-2.51) | **1.85 (1.04-3.28) *** |
| High waist circumference (NCEP ATP-III criteria) g | 1.00 (Reference) | 0.20 (0.04-1.11) | 0.83 (0.35-2.00) | 1.00 (Reference) | 0.80 (0.36-1.78) | 0.90 (0.49-1.66) |
| High waist circumference (Consensus criteria) g | 1.00 (Reference) | **0.18 (0.04-0.88) *** | 1.53 (0.40-5.79) | 1.00 (Reference) | 0.49 (0.20-1.19) | 0.72 (0.35-1.50) |
| Mets (NCEP ATP-III criteria) f | 1.00 (Reference) | 0.50 (0.13-1.88) | 0.79 (0.37-1.68) | 1.00 (Reference) | 0.75 (0.36-1.58) | 1.07 (0.62-1.84) |
| Mets (Consensus criteria) f | 1.00 (Reference) | 0.58 (0.16-2.13) | 1.48 (0.65-3.36) | 1.00 (Reference) | 0.87 (0.42-1.80) | 1.06 (0.61-1.85) |
| **p-value <0.05* |  |  |  |  |  |  |
| *f The logistic regression model adjusted for gender, education, marital status, physical activity, alcohol usage, tobacco usage and cardiac illness* | | | | | | |
| *g Each component of Mets was adjusted for other components along with gender, marital status, physical activity, alcohol usage, tobacco usage and cardiac illness* | | | | | | |
